# Leverage Real-world Longitudinal Data in Large Clinical Research Networks for Alzheimer’s Disease and Related Dementia (ADRD)

**DOI:** 10.1101/2020.08.03.20167619

**Authors:** Rui Duan, Zhaoyi Chen, Jiayi Tong, Chongliang Luo, Tianchen Lyu, Cui Tao, Demetrius Maraganore, Jiang Bian, Yong Chen

**Affiliations:** Department of Biostatistics, Harvard T.H. Chan School of Public Health, Boston, MA, USA; Department of Epidemiology, College of Medicine & College of Public Health and Health Professions, University of Florida, Gainesville, FL, USA; Department of Biostatistics, Epidemiology and Informatics, Perelman School of Medicine, The University of Pennsylvania, Philadelphia, PA, USA; Department of Health Outcomes and Biomedical Informatics, College of Medicine, University of Florida, Gainesville, FL, USA; School of Biomedical Informatics, The University of Texas Health Science Center at Houston, Houston, TX, USA; Department of Neurology, College of Medicine, University of Florida, Gainesville, FL, USA

## Abstract

With vast amounts of patients’ medical information, electronic health records (EHRs) are becoming one of the most important data sources in biomedical and health care research. Effectively integrating data from multiple clinical sites can help provide more generalized real-world evidence that is clinically meaningful. To analyze the clinical data from multiple sites, distributed algorithms are developed to protect patient privacy without sharing individual-level medical information. In this paper, we applied the One-shot Distributed Algorithm for Cox proportional hazard model (ODAC) to the longitudinal data from the OneFlorida Clinical Research Consortium to demonstrate the feasibility of implementing the distributed algorithms in large research networks. We studied the associations between the clinical risk factors and Alzheimer’s disease and related dementia (ADRD) onsets to advance clinical research on our understanding of the complex risk factors of ADRD and ultimately improve the care of ADRD patients.

## Introduction

Over the last few decades, there has been increased adoption of electronic health records (EHR) systems in the United States (US). Containing rich medical information, EHR data can potentially improve the efficiency and effectiveness of health care and biomedical research^1,2^. EHR data are considered as real-world data (RWD) that are collected outside of traditional clinical research settings (e.g., randomized controlled trials). The U.S. Food and Drug Administration (FDA) recently launched the real-world evidence (RWE) program to encourage the use of RWD such as EHRs, administrative claims, and billing data among others, to support the development, evaluation, and monitoring of drug products^3^, where methods that can reliably measure disease progression and the impact of the drugs are urgently needed. The detailed temporal information contained in EHR, including conditions and diagnoses, procedures, medications, laboratory test results, disease status, and treatment outcomes, create opportunities for analyzing time-to-event outcomes, which can provide a better understanding of the disease progression and impact of the treatments or risk factors on the timing of outcomes. Among many methods developed for analyzing time-to-event data^4^, the Cox proportional hazards model^3^ is one of the commonly used methods and has been widely applied in biomedical research.

To better utilize EHR data collected at different clinical sites, increasing numbers of data consortia were founded over the last few decades^5–7^. The national Patient-Centered Clinical Research Network (PCORnet), which is funded by Patient-Centered Outcomes Research Institute (PCORI), covers more than 100 million patients all over the United States^6^. As one of the nation’s 9 clinical data research networks (CDRNs), the OneFlorida Clinical Research Consortium (OneFlorida CRC) was designed to accelerate the translation of promising research findings into improved patient care, with a focus on comparative effectiveness research, programmatic clinical trials, and patient-centered outcomes studies. The centerpiece of the OneFlorida network is its Data Trust—a repository of statewide health care data containing robust longitudinal and linked patient-level RWD of around 15 million (>50%) Floridians, including data from Medicaid claims, cancer registries, vital statistics, and EHRs from its clinical partners^8^. Currently, there are 12 various types of healthcare organizations contributing to the OneFlorida data repository: 1) four academic health centers (i.e., University of Florida Health, University of Miami Health System, Florida State University and regional campus practice partners, and University of South Florida Health), 2) seven healthcare systems including Tallahassee Memorial Healthcare (affiliated with Florida State University), Orlando Health, Adventist Health, Nicklaus Children’s Hospital, Bond Community Health, Capital Health Plan, and Tampa General Hospital, and 3) CommunityHealth IT— a rural health network in Florida. Covering all 67 Florida counties^9^, the OneFlorida CRC provides care for more than 50% of Floridians through 4,100 physicians, 914 clinical practices, and 22 hospitals, leading to the increasing amount of longitudinal and robust patient-level records of around 15 million Floridians and over 561.1 million encounters,1.16 billion diagnoses, 1 billion prescribing records, and 1.44 billion procedures as of December 2019.

The large-scale clinical data networks like the OneFlorida CRC allow researchers to study the diverse range of risk factors, from clinical characteristics to social determinants of health, associated with the fatal degenerative diseases: Alzheimer’s disease (AD) and AD-related dementia (ADRD). Alzheimer’s disease (AD) is the most common cause of dementia. In 2019, 5.8 million Americans will live with AD, among which 97% are aged ≥ 65 and 81% are ≥75. By 2050, people living with AD in the US may grow to 13.8 million, fueled by the aging baby boomers^10^. In 2017, 121,404 deaths from AD were recorded, making AD the 6th leading cause of death and the 5th leading cause of death among Americans aged ≥ 65^11^. The progression of AD/ADRD usually starts with normal cognition during the preclinical period, slowly advancing to mild cognitive impairment (MCI) and then gradually progresses to mild and moderate AD and eventually severe AD. It usually takes an individual with MCI 7 years to progress to mild AD, but some individuals may experience a rapid progression which took significantly less time to develop into AD^12^. Therefore, modeling the disease progression in an at-risk population is important.

Combining RWD from multiple clinical sites can provide a larger sample size, which can lead to more generalizable findings, and the ability to better evaluate rare risk factors of ADRD. Nevertheless, directly sharing individual patient-level data can be challenging because of the privacy concerns over protected patient health information^14^. The state-of-the-art method for multicenter study, without sharing patient-level information, is the meta-analysis, where each site fits separate analysis and all local estimates are synthesized using a weighted average^15^. In addition to meta-analysis, several distributed learning algorithms, in which only aggregate information is allowed to be shared across institutions, have been developed to overcome the privacy issue in a clinical research network^16–19^. Among the existing methods, Duan et al^20^. proposed a privacy-preserving and communication-efficient distributed algorithm, named ODAC, to fit the multi-center Cox proportional hazards model. Utilizing a surrogate likelihood approach^21^ and without iterative communication across the sites, the ODAC algorithm allows efficient and accurate identification of the risk factors associated with the time-to-event outcome of interest^20^. This algorithm is shown to achieve high accuracy in the sense that the results of ODAC is close to the pooled analysis in which a Cox model is fitted on the combined dataset.

In this paper, with the multicenter RWD data from OneFlorida, we evaluated the ODAC algorithm using ADRD as a use case. By studying the associations between ADRD and several clinical risk factors, 7 significant risk factors were identified by ODAC, and the 7 risk factors were consistent with previous findings on ARDR. In addition, compared with the commonly used meta-analysis, ODAC provided estimates of the effect sizes with smaller bias, leading to the conclusion that ODAC is a reliable and efficient, privacy-preserving distributed learning algorithm extremely suitable for working with real-world data from multicenter clinical research networks.

## Materials and Methods

### Data Source and Study Population

The OneFlorida data is a HIPAA (i.e., Health Insurance Portability and Accountability Act) limited data set (i.e., dates are not shifted; and 9-digit zip codes of patients’ residencies are available, where other 16 types of patient identifiers were removed) that contains detailed patient and clinical variables, including demographics, encounters, diagnoses, procedures, vitals, medications, and labs, following the PCORnet Common Data Model (CDM)^6^. The OneFlorida data undergo rigorous quality checks at its data coordinating center (i.e., University of Florida [UF]), and a privacy-preserving record linkage process is used to deduplicate records of same patients coming from different health care systems within the network^22^.

Based on the OneFlorida data, individuals who were 65 years of age, and had no ADRD diagnosis before 2014/03/01 (i.e., the index date) were considered as “at-risk population” and were included in our analysis. We chose 2014/03/01 as the baseline to ensure a 5-year follow-up period for each individual. The outcome of interest is the time to the first diagnosis of ADRD. Conditions that were considered as ADRD include mild cognitive impairment (ICD-9: 331.81, 294.9; ICD-10: G31.83, F09), Alzheimer’s disease (ICD-9: 331.0; ICD-10: G30.0, G30.1, G30.8, G30.9), vascular dementia (ICD-9: 290.40, 290.41; ICD-10: F01.50, F01.51), Lewy body dementia (ICD-9: 331.82; ICD-10: G31.83), Frontotemporal Dementia (ICD-9: 331.19; ICD-10: G31.09), and primary progressive aphasia (ICD-9: 331.11; ICD-10: G31.01). A total of 178,251 patients who were at risk at baseline were identified in OneFlorida. After excluding patients with missing data, a total of 16,456 individuals were included in our final analysis. **Table 1** shows the prevalence of the included population across the different OneFlorida clinical sites.

**Table 1.** Breakdown of the study populations by site.

| Site | ADRD | At-risk population | prevalence |
| --- | --- | --- | --- |
| Site 1 | 128 | 3,292 | 5.34% |
| Site 2 | 10 | 983 | 1.02% |
| Site 3 | 711 | 12,181 | 5.84% |
| **All sources** | **849** | **16,456** | **5.16%** |

### Risk Factors

We identified a set of risk factors from the literature and extracted the factors from patients’ medical records in OneFlorida. All records before the index date (2014/03/01) were taken into consideration in this analysis. Factors such as demographic variables (age, race, gender, and insurance type), vital signs (body mass index [BMI], lipid panel), smoking status, selected clinical diagnoses, and medications were included. Since the laboratory test results (e.g., complete blood count) had high frequencies of missing values (>50% in the total study population), these factors were removed. Besides, the clinical diagnoses that were made in <1% of the total study population were removed to minimize potential bias introduced by the small sample size. Patients who had missing values in any risk factors were removed as the current ODAC algorithm is unable to handle missing values. A total of 12 risk factors/predictors were included in the analysis. In **Table 2**, we present the summary statistics of these predictors. From the table, we observed that the patients who were diagnosed with ADRD were older, more males, and tended to have Medicare insurance. In addition, ADRD patients were more likely to have more comorbidities and higher statin use.

**Table 2.** Characteristics of included risk factors in the cohort.

|  | **Control**<br>**(n=15607)** | **Case**<br>**(n=849)** | **Overall**<br>**(n=16456)** |
| --- | --- | --- | --- |
| **Baseline AGE** |  |  |  |
| Mean (SD) | 73.9 (5.94) | 76.5 (6.54) | 74.0 (6.00) |
| **Gender** |  |  |  |
| Female | 8981 (57.5%) | 474 (55.8%) | 9455 (57.5%) |
| Male | 6626 (42.5%) | 375 (44.2%) | 7001 (42.5%) |
| **RACE_ETHNICITY** |  |  |  |
| Hispanic | 277 (1.8%) | 20 (2.4%) | 297 (1.8%) |
| NHB* | 2324 (14.9%) | 141 (16.6%) | 2465 (15.0%) |
| NHW* | 12588 (80.7%) | 667 (78.6%) | 13255 (80.5%) |
| Other | 418 (2.7%) | 21 (2.5%) | 439 (2.7%) |
| **insurance_type** |  |  |  |
| MEDICAID | 140 (0.9%) | 4 (0.5%) | 144 (0.9%) |
| MEDICARE | 11568 (74.1%) | 694 (81.7%) | 12262 (74.5%) |
| No payment* | 48 (0.3%) | 2 (0.2%) | 50 (0.3%) |
| Other | 470 (3.0%) | 3 (0.4%) | 473 (2.9%) |
| OtherGOV | 161 (1.0%) | 19 (2.2%) | 180 (1.1%) |
| PRIVATE | 3220 (20.6%) | 127 (15.0%) | 3347 (20.3%) |
| **BMI** |  |  |  |
| Mean (SD) | 28.9 (6.18) | 27.9 (6.75) | 38.5 (889) |
| **SMOKING** |  |  |  |
| Current Smoker | 1000 (6.4%) | 46 (5.4%) | 1046 (6.4%) |
| Former smoker | 6301 (40.4%) | 337 (39.7%) | 6638 (40.3%) |
| Never smoker | 8306 (53.2%) | 466 (54.9%) | 8772 (53.3%) |
| **depression** | 1837 (11.8%) | 175 (20.6%) | 2012 (12.2%) |
| **anxiety** | 1859 (11.9%) | 145 (17.1%) | 2004 (12.2%) |
| **sleep_disorder** | 1578 (10.1%) | 123 (14.5%) | 1701 (10.3%) |
| **hypertension** | 10208 (65.4%) | 600 (70.7%) | 10808 (65.7%) |
| **diabetes** | 4081 (26.1%) | 254 (29.9%) | 4335 (26.3%) |
| **heart_disease** | 1701 (10.9%) | 136 (16.0%) | 1837 (11.2%) |
| **statin** | 4646 (29.8%) | 333 (39.2%) | 4979 (30.3%) |

\*NHB stands for non-Hispanic black; NHW stands for non-Hispanic White; *No payment* includes self-pay, nor charge, refusal to pay/bad debt, hill burton free care, research/donor, and other.

### Statistical Analysis

To help introducing the ODAC algorithm, we first define some basic notation. Suppose we have sites in the clinical network, where the j-th site has *n*_*j*_ samples, with 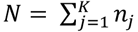. For the i-th subject in the j-th site, we observe {*T*_*ij*_,*δ*_*ij*_,*x*_*ij*_}where *x*_*ij*_ is a vector denoting risk factors, *T*_*ij*_ is the time-to-event for the outcome of interest, and *δ*_*ij*_ is the event indicator with *δ*_*ij*_ = 1indicating an event, and *δ*_*ij*_ = 0 indicating an censored. We apply the distributed algorithm ODAC developed in Duan et al.^20^, and the detailed algorithm is shown below with the definition of each quantity in **Table 3**. The main idea of the algorithm is to fit Cox model at each site first and combine the results through a fixed-effect meta-analysis to obtain initial values for regression parameters, and then apply the surrogate likelihood approach proposed by Jordan et al.^21^ which requires each site to calculate summary-level quantities for calculating the first and second-order derivatives of the combined log-partial likelihood function. A surrogate likelihood can serve as a good proxy of the combined log-partial likelihood function, and the final estimator is obtained by maximizing the surrogate function. **Figure 1** provides a schematic illustration of the above algorithm,and we refer to Duan et al.^20^ for more technical details and properties of the ODAC method. In addition to ODAC, we also applied the meta-analysis and the pooled analysis to the multicenter EHR data we extracted from OneFlorida. *NHB stands for non-Hispanic black; NHW stands for non-Hispanic White; *No payment* includes self-pay, nor charge, refusal to pay/bad debt, hill burton free care, research/donor, and other.

**Table 3.** Descriptions and definitions of quantities for ODAC.

| Quantity | Description | Definition |
| --- | --- | --- |
| $R_j(t)$ | Risk set at time $t$ in the $j$-th site | $R_j(t) = \{i; T_{ij} \geq t\}$ |
| $L_j(\beta)$ | Log partial likelihood function at $j$-th site | $\frac{1}{n_j} \sum_{i=1}^{n_j} \delta_{ij} \log \frac{\exp(\beta^T x_{ij})}{\sum_{s \in R_j(T_{ij})} \exp(\beta^T x_{sj})}$ |
| $L(\beta)$ | Combined log partial likelihood function | $L(\beta) = \sum_{j=1}^K \frac{n_j}{N} L_j(\beta)$ |
| $\hat{\beta}_j$ | Local estimate at $j$-th site | $\text{argmax}_{\beta} L_j(\beta)$ |
| $\hat{V}_j$ | Variance of $\hat{\beta}_j$ | $-\{n_j \nabla^2 L_j(\hat{\beta}_j)\}^{-1}$ |
| $\bar{\beta}$ | Initial value for $\beta$ | $(\sum_{j=1}^K \hat{V}_j^{-1})^{-1} \sum_{j=1}^K \hat{V}_j^{-1} \hat{\beta}_j$ |
| $\mathcal{T}$ | Collection of all time points | $\{T_{ij}; \delta_{ij} = 1\}$ |
| $U_j(t)$ | Intermediate results | $\sum_{i \in R_k(t)} \exp(\bar{\beta}^T x_{ik})$ |
| $W_j(t_s)$ | Intermediate results | $\sum_{i \in R_k(t)} \exp(\bar{\beta}^T x_{ik}) x_{ik}$ |
| $Z_j(t_s)$ | Intermediate results | $\sum_{i \in R_k(t)} \exp(\bar{\beta}^T x_{ik}) x_{ik} x_{ik}^T$ |
| $\tilde{L}_j(\beta)$ | Surrogate likelihood function at $j$-th site | $L_j(\beta) + \langle \nabla L(\bar{\beta}) - \nabla L_j(\bar{\beta}), \beta \rangle + \frac{1}{2} (\beta - \bar{\beta})^T \{\nabla^2 L(\bar{\beta}) - \nabla^2 L_j(\bar{\beta})\} (\beta - \bar{\beta})$ |
| $\tilde{\beta}_k$ | Surrogate estimator in $j$-th site | $\text{argmax}_{\beta} \tilde{L}_j(\beta)$ |
| $\tilde{V}_k$ | Estimated variance of $\tilde{\beta}_k$ | $\tilde{V}_j = \frac{1}{N} \{-\nabla^2 L_j(\tilde{\beta}_j)\}^{-1}$ |
| $\tilde{\beta}$ | ODAC estimator | $(\sum_{j=1}^K \tilde{V}_j^{-1})^{-1} \sum_{j=1}^K \tilde{V}_j^{-1} \tilde{\beta}_j$ |

**Figure 1.**
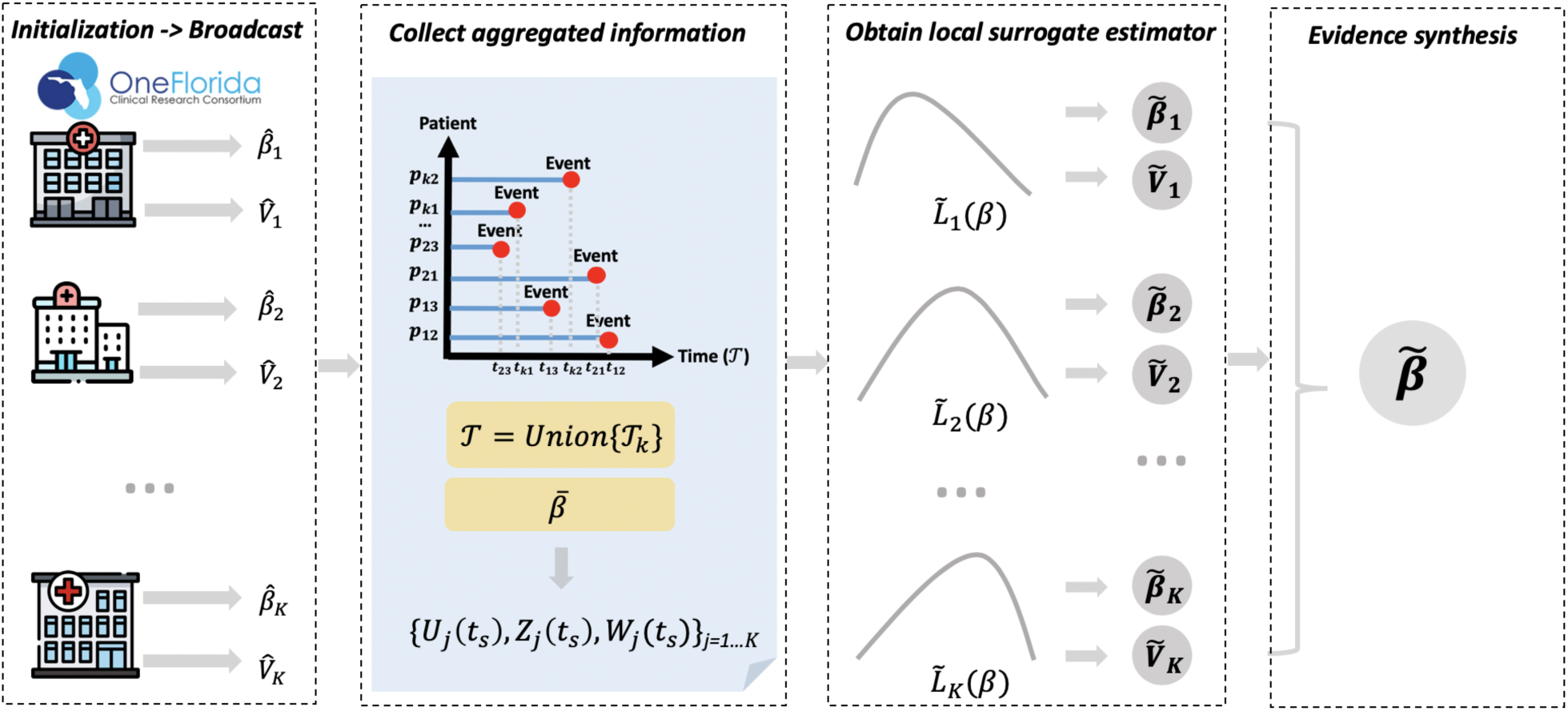
Illustration of the ODAC algorithm. The first step is initialization, in which each site reports the local estimate of the log hazard ratio 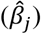 and the variance 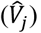 where j = 1, …,K. Then with these initial values broadcasted across the sites, the average of all local estimates and the union of all event times can be obtained. With these results, each site calculates and shares the intermediate results {*U*_*j*_ (*t*_*s*_), *W* _*j*_(*t*_*s*_)}. Next, within each site, the intermediate results and individual patient-level data are used to construct the local surrogate likelihood function. By optimizing the surrogate likelihood function, we obtain the local surrogate estimates. The final step is to synthesize the surrogate estimates from all the sites.

#### Algorithm ODAC

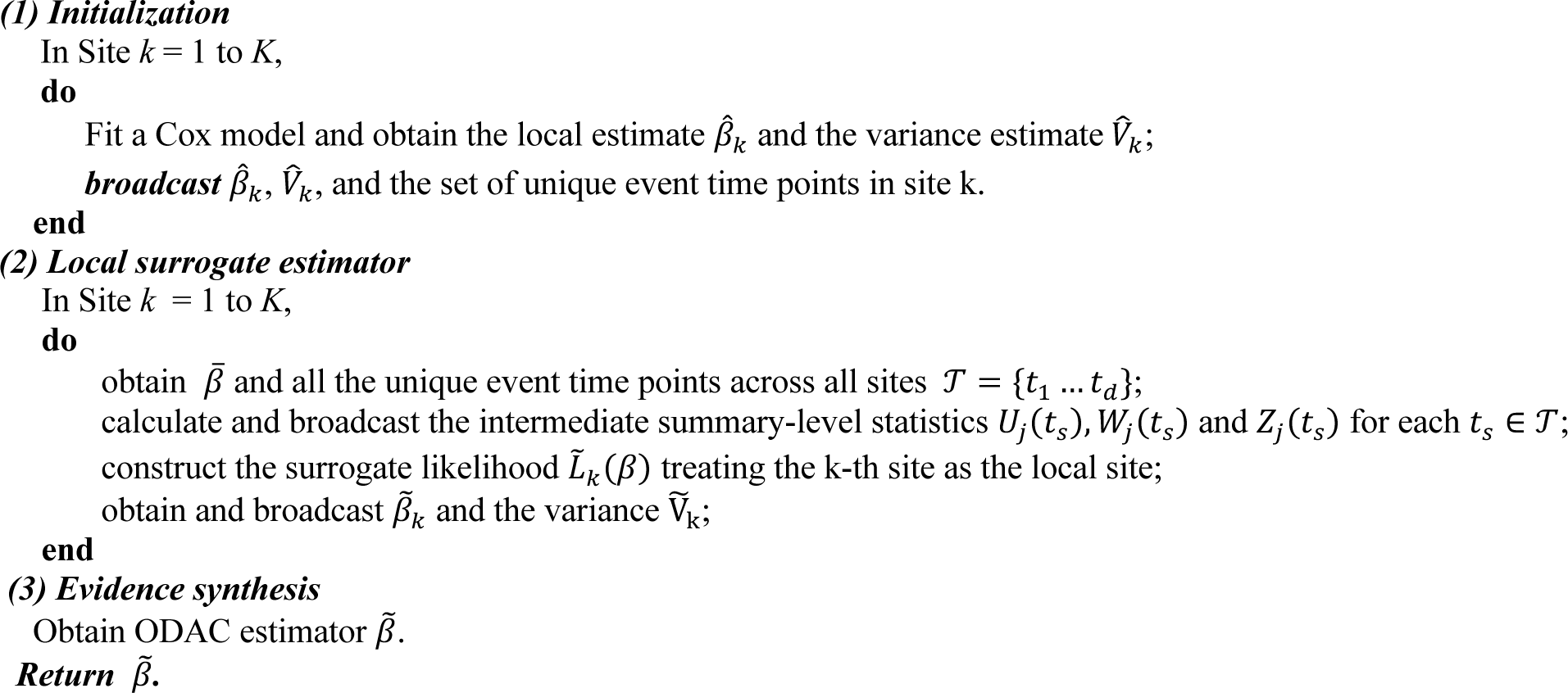

## Results

**Figure 2** shows the estimated log hazard ratio and 95% confidence interval for each risk factor using the three different approaches: the pooled analysis, meta-analysis, and ODAC. The pooled analysis is considered as the gold standard, since it essentially requires sharing all the patient-level data across sites. From the plot, we observe that ODAC provides more accurate estimates for the associations between outcome and exposures than meta-analysis, in the sense that the bias to the gold standard pooled estimator is smaller. To be more specific, ODAC estimates are closer to the pooled estimates for 11 out of the 13 risk factors we considered in the Cox regression model. The relative bias for the rest of two risk factors are below 6%. The bias for meta-analysis is up to 185% For risk factors such as BMI and statin, the conclusion regarding the significance of the risk factor from meta-analysis is not consistent with the pooled analysis, while ODAC provides the same conclusion as the pooled analysis. The confidence intervals of ODAC is observed to be slightly larger than the other two methods, which might be caused by the heterogeneity of the data across sites.

**Figure 2.**
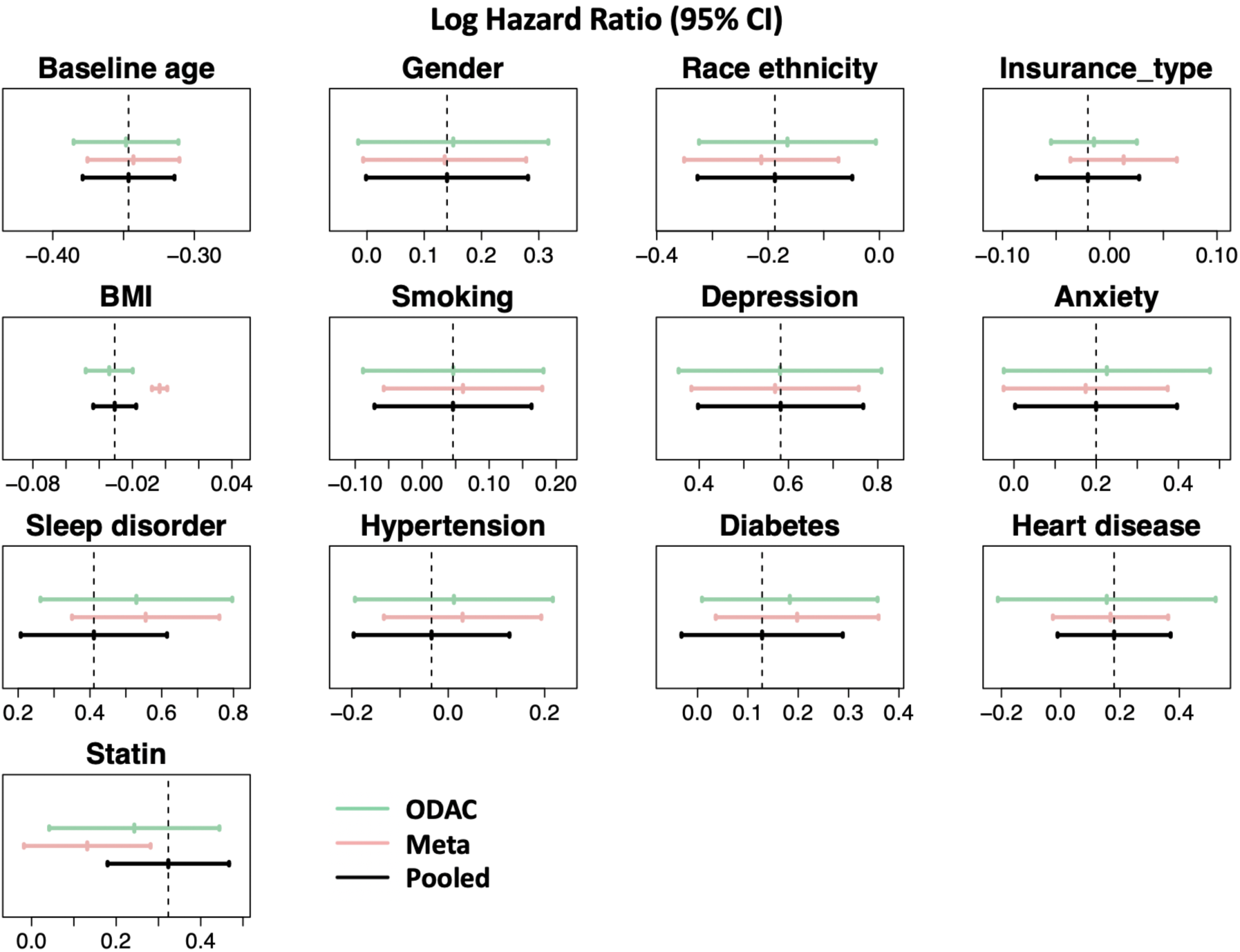
displays the estimated log hazard ratio and 95% confidence interval for each risk factor using the three methods: ODAC (green), meta-analysis (light pink), and pooled estimate (black).

From the analysis, we identified 7 significant risk factors: anxiety, BMI, sleep disorder, statin, age, depression, and race. Most of our findings are consistent with previous reports on risk factors of ADRD. For example, demographic variables such as female, being non-Hispanic Blacks were more likely to have higher risks of ADRD. Cardiovascular conditions, including hypertension, heart diseases could also increase a person’s risk of having ADRD^23–25^. Mental health conditions, such as anxiety, depression, or sleep disorder, have also been demonstrated to be associated with ADRD, as patients who have ADRD tend to have symptoms that lead to decline in mental functions^23,26^. Interestingly, statins use were shown to be positively associated with the occurrence of ADRD in our data, which is contradicted to previous reports^27^ that statin may be protective against the progression of AD, this is possibly due to the heterogeneity in the ADRD population^28,29^ and potential misclassifications of how medication prescriptions are recorded in the data.

## Discussion and conclusion

In this paper, with the longitudinal EHR data from the OneFlorida CRC, we evaluated the empirical performance of a privacy-preserving, communication-efficient distributed learning algorithm—the ODAC algorithm. We studied the associations between ADRD and 13 risk factors and identified 7 significant factors that are consistent with the previous findings on ARDR. In the evaluation, other than applying ODAC, we also conducted the meta-analysis, which is currently the most popular method for multicenter analysis. From the results, we observed that the ODAC algorithm is able to provide improved estimation of effect sizes for the risk factors compared with the meta-analysis. In ODAC, the strategy that each clinical site serves as the local site reduces the potential impact of one local site on the final estimate. Different from the other existing distributed algorithms, the ODAC is a non-iterative algorithm by constructing the surrogate likelihood function without sharing patient-level information. In a nutshell, ODAC is a practical, robust, efficient, and accurate algorithm for modeling time-to-event outcomes.

Even though the pathophysiology of AD and ADRD is not well understood, there is evidence indicating the heterogeneity in AD as well as the heterogeneity in progression to AD through different intermediate disease stages. Identification of the factors contributing to different progression pathways from MCI to AD are crucial for clinical prognostication and risk stratification to guide counseling and selection of potential treatments. RWD from large clinical networks similar to OneFlorida provide the golden opportunity to examine the heterogeneity of AD. Distributed learning algorithms like ODAC, thus, become critical to leverage these RWD to generate RWE. Risk models built with ODAC can help us identify critical factors for both the primary prevention (i.e., from non-AD to AD) and secondary prevention (i.e., from MCI to AD) of AD, which ultimately lead to better care for ADRD patients.

In this study, we only used the structured data from the OneFlorida network, where some other important risk factors, especially social determinants of health (SDoH) were not readily available and thus not included in our analysis. On the other hand, clinical narratives^30^ in EHR contain more detailed patient information, including SDoH. Further, we only were able to model ADRD onset as the outcome. To accurately study the progression of ADRD, we would need to be able to extract and model other intermediate outcomes such as neuropsychological tests (e.g., Mini-Mental State Examination and Severe Impairment Battery), that are not typically captured in structured EHR either. In future studies, advanced natural language processing (NLP) methods can be leveraged to extract additional risk factors and neuropsychological test results from clinical narratives.

In the future, the ODAC algorithm can be extended in several aspects. First, the distributed algorithms to integrate and study other types of outcomes can be considered, for example, count data and longitudinal outcomes. Secondly, ODAC is based on the fact that we treat the pooled analysis as the gold standard method, which essentially requires data to be homogenous distributed across sites. However, the effect sizes as well as the baseline hazard function at each site can be different in practice^31^. To efficiently synthesize evidence from heterogeneous clinical sites in large CRN, we are planning to extend the current distributed algorithms by allowing site-specific hazard function and effect sizes in the future. Thirdly, we will work on developing distributed algorithm to handle time-varying covariates or time-varying coefficients in survival analysis. Lastly, we have been developing an open-source software to implement the ODAC algorithm within CRN to facilitate data integration and nationwide comparative effectiveness studies.

## Data Availability

N/A

## Acknowledgement

This work was supported in part by NIH grants 1R01AI130460, 1R01LM012607, R01CA246418, R21AG061431 and UL1TR001427, and PCORI grants ME-2018C3-14754. The content is solely the responsibility of the authors and does not necessarily represent the official views of the NIH and PCORI.

